# Subtypes of Relapsing-Remitting Multiple Sclerosis Identified by Network Analysis

**DOI:** 10.1101/2022.11.16.22282420

**Authors:** Quentin Howlett-Prieto, Chelsea Oommen, Michael D. Carrithers, Donald C. Wunsch, Daniel B. Hier

**Affiliations:** Department of Neurology and Rehabilitation, University of Illinois at Chicago, Chicago IL USA; Department of Electrical and Computer Engineering, Missouri University of Science and Technology, Rolla MO USA

**Keywords:** multiple sclerosis, phenotype, network analysis, communities, modularity

## Abstract

The objective of this study was to use network analysis to identify subtypes of relapsing-remitting multiple sclerosis subjects based on their cumulative signs and symptoms. We reviewed the electronic medical records of 120 subjects with relapsing-remitting multiple sclerosis and recorded signs and symptoms. Signs and symptoms were mapped to a neuro-ontology and then collapsed into 16 superclasses by subsumption and normalized. Bipartite (subject-feature) and unipartite (subject-subject) network graphs were created using Gephi. Degree and weighted degree were calculated for each node. Graphs were partitioned into communities using the modularity score. Feature maps were used to visualize differences in features by the community. Network analysis of the unipartite graph yielded a higher modularity score (0.49) than the bipartite graph (0.247). Network analysis can partition multiple sclerosis subjects into communities based on signs and symptoms. Communities of subjects with predominant motor, sensory, pain, fatigue, cognitive, behavior, and fatigue features were found. Larger datasets and additional partitioning algorithms are needed to confirm these results and elucidate their clinical significance.

## INTRODUCTION

Multiple sclerosis (MS) is one of several immune-mediated demyelinating diseases of the central nervous system that includes transverse myelitis, optic neuritis, neuromyelitis optical, acute disseminated encephalomyelitis, and acute hemorrhagic leukoencephalopathy [1]. MS has traditionally been divided into four phenotypes based on clinical course that include relapsing-remitting multiple sclerosis (RRMS), primary progressive multiple sclerosis (PPMS), secondary progressive multiple sclerosis (SPMS), and relapsing progressive multiple sclerosis (RPMS) [2]. In 2013, the criteria for MS phenotypes were revised to delete RPMS [3, 4, 5, 6]. More recently, there has been a trend to classify MS cases by activity (active or inactive) or by phase (relapsing or progressive) [4, 5, 7]. Furthermore, the clinical presentation of MS may differ by race [8, 9, 10]. Studies have suggested more cognitive impairment [8], a higher prevalence of optico-spinal and transverse myelitis presentations, [9], and a faster rate of brain and retinal atrophy in African American patients with MS [10].

Another potential way of subtyping neurologic diseases is by the burden of neurological signs and symptoms. This approach has been useful in identifying subtypes of frontotemporal dementia (behavioral and aphasic variants), multiple system atrophy (cerebellar and Parkinsonian variants), amyotrophic lateral sclerosis (typical, primary lateral sclerosis, progressive bulbar atrophy, progressive muscular atrophy, and other variants), and Guillain-Barre syndrome (typical, Miller-Fisher variant, and axonal variants) [11, 12, 13, 14]. Poser [15] emphasized that MS may have a variable onset with symptoms as diverse as optic neuritis, facial pain, hemifacial spasm, Lhermitte sign, transverse myelitis, limb weakness, limb numbness, urinary retention, dysmetria, intention tremor, incoordination, dysarthria, hearing loss, color blindness, gait disturbance, and diplopia. Subtypes of multiple sclerosis based on clinical presentation (signs and symptoms) are recognized [16, 17, 18, 19]. MS patients can present with different predominant symptoms that may emphasize tremor [20], ataxia [21], visual symptoms [22, 23], sensory symptoms including numbness and paresthesias [24, 25, 26, 27], pyramidal tract findings such as weakness, hyperreflexia, spasticity, and hypertonia [28, 29, 30], or spinal cord presentations that include paraparesis, sphincter dysfunction, and sensory levels [31, 32]. Some patients present with cognitive impairment [33, 34], dysarthria [35], dysautonomia [36], depression and psychiatric disturbances [37], imbalance [38] or fatigue [39]. It is of interest whether identifiable subtypes of MS based on predominant clinical presentation (sensory, cerebellar, motor, visual, cognitive, fatigue, etc.) can be reliably identified. Clinical subtypes could differ with regard to biomarker profile, prognosis, course, and response to treatment. The network analysis of signs and symptoms could assist in identifying clinically significant subtypes of MS.

A network (also called a graph) is an assembly of nodes that are interconnected by edges [40]. When all connected nodes come from the same class, the graph is called unipartite. When each node is connected to a node of a second class, the graph is called bipartite. Networks can be created from large medical datasets and analyzed without a priori knowledge of how the nodes connect [41]. Algorithms can partition networks into communities of interconnected nodes (also called clusters) [42]. Barabási [43] defines a community as “a locally dense connected subgraph in a network (page 325)” and that “…we expect nodes that belong to a community to have a higher probability of linking to other members of that community than to nodes that do not belong to the same community “ Many partitioning algorithms depend upon the maximization of a metric called modularity which measures how well each community is separated from other communities.

Network analysis has proven useful in visualizing the complex relationships between the phenotypes, genes, proteins, and metabolic pathways that underlie human diseases (Table 1) [44, 45]. Network analysis has been applied to identifying potential genetic causes of autism [46] as well as the clustering of autism subjects by phenotype [47, 48]. Network analysis has been used to identify genes that may govern MS susceptibility [49] as well as brain areas that may undergo disconnection in MS [50, 51, 52, 53].

**Table 1.** Some examples of the application of network analysis to problems in human disease. This list is representative and not definitive. In general, network analysis begins with identifying nodes and edges. When nodes are connected to nodes of the same class, the graph is unipartite. When nodes are connected to a node of a second class, the graphs are bipartite. Edges connect the nodes.

| Graph | Domain | node | node | edge | Citation |
| --- | --- | --- | --- | --- | --- |
| unipartite | phenotypes | phenotype | phenotype | similarity | [55] |
| unipartite | metabolism | metabolic pathway | metabolic pathway | interaction | [56] |
| unipartite | diseases | disease | disease | phenotype | [57] |
| unipartite | proteins | protein | protein | interaction | [57] |
| bipartite | proteins | disease | protein | disease-gene | [57] |
| bipartite | heart disease | heart disease | disease | co-morbidity | [58] |
| unipartite | autism | gene | gene | co-expression | [59] |
| unipartite | autism | gene | gene | co-expression | [46] |
| bipartite | autism | autism | disease | association | [60] |
| unipartite | autism | subject | subject | phenotype | [48, 47] |
| unipartite | brain disease | brain area | brain area | connectivity | [61] |
| unipartite | interactome | disease | disease | metabolism | [62] |
| unipartite | MS | brain area | brain area | connectivity | [50, 51, 52, 53] |
| unipartite | MS | gene | gene | susceptibility | [49] |
| unipartite | MS | MS patient | MS patient | sign/symptom | † |
| bipartite | MS | MS patient | phenotype | sign/symptom | † |

† This paper

### Proposed Approach

Our goal was to determine if clinical subtypes of RRMS based signs and symptoms could be identified. We found 244 unique neurologic signs and symptoms in a cohort of subjects with relapsing-remitting MS and collapsed them into sixteen superclasses. The count of signs and symptoms in each superclass was normalized for each subject. A bipartite graph was created in Gephi, with each subject node connected to each of the sixteen superclass nodes by an edge proportional to the normalized count of signs and symptoms. Distances between subjects were calculated by the cosine similarity of their signs and symptoms. A unipartite graph was created in Gephi where the nodes were subjects, and the edges were subject similarities. The unipartite and bipartite graphs were partitioned into communities based on the Louvain algorithm [54]. We used modularity scores to evaluate the quality of the partition and heat maps to evaluate the composition of the communities.

## METHODS

### Subjects

One hundred and twenty MS subjects followed at the University of Illinois-Neuroscience Center were enrolled in the University of Illinois at Chicago (UIC) Neuroimmunology Biobank between August 2018 and March 2020 (mean age 42.7 *±* 12.8 years, 73% female, 27% male, 58% Black, 42% White). The Biobank is approved by the Institutional Review Board (IRB) of the University of Illinois College of Medicine. All subjects provided informed written consent at enrollment. Subjects were between 18-80 years old and had a diagnosis of RRMS based on the 2017 McDonald criteria [63]. Subjects had been recruited for a study of blood biomarkers in MS where RRMS was an inclusion criterion, and progressive MS was an exclusion criterion. Seven subjects with normal neurological examinations were excluded from the analysis leaving a final study sample of 113 subjects.

### Neuro-phenotyping

The neurological progress notes from the electronic health record of all subjects were reviewed, and neurological signs and symptoms were recorded [64]. For the purposes of this study, the cumulative signs and symptoms (both active and resolved) of each subject were recorded. Signs and symptoms were mapped to concepts in a neuro-ontology that had 1600 possible concepts [65]. Subjects had 13.2 ∓ 9.2 signs and symptoms (mean ∓ standard deviation). The 113 subjects had 1453 total signs and symptoms (244 unique signs and symptoms). We used the hierarchy of the neuro-ontology [66] to collapse by subsumption the signs and symptoms into 16 superclasses that included *behavior, cognitive, cranial nerve, eye movement, fatigue, gait, hyperreflexia, hypertonia, incoordination, pain, sensory, speech, sphincter, tremor, vision*, and *weakness* Each subject was represented as a 17-dimension vector. The first element of the vector was the case identification label, and the subsequent sixteen elements were the count of signs and symptoms for each of the sixteen superclasses. The final dataset was a 113 × 17 element array with rows as instances (subjects) and columns as features (superclasses of signs and symptoms). A normalized array was created by normalizing each feature over the interval [0,1] using the *continuize* widget in Orange 3.32.0 [67]. For the availability of python programs and datasets, see the Data Availability statement.

### Network analysis, distance metrics, feature maps

Network analyses were performed on normalized 113 × 17 data arrays [67, 68, 69]. Bipartite networks were visualized in Gephi 0.9.7 using a variety of layouts, with the final analysis using the Force Atlas layout with a repulsion force of 10,000. Visual inspection showed Force Atlas to have the optimal spacing of nodes and clarity of visualization. The bipartite network contained nodes of subjects and features (signs and symptoms) as nodes with a magnitude of the edges connecting subjects to features equal to the normalized feature score for each subject. In the bipartite networks, there were no direct subject-subject or feature-feature edges. Node sizes were proportional to the average weighted degree of each node. Communities were named based on their predominant features. Nodes were colored by their community membership, and colors were used consistently across graphs based on feature predominance. Edge widths were proportional to edge weight for the bipartite graphs.

The unipartite networks were based on distances between subjects derived from the feature vectors for each subject. Distances were calculated in Orange using the *distances* widget for Pearson, Euclidean, and cosine distances. Visual inspection of the network graphs showed that the cosine-based graphs were superior to those based on the Pearson or Euclidean distances. Only the cosine distances were retained for further analysis [70]. For the unipartite graphs, all nodes were subjects, and the edges were subject similarity based on the cosine distances. Node size was proportional to degree (number of edges for each node). Edge width was fixed.

Gephi was used to partition the unipartite and bipartite networks into communities based on the Louvain algorithm [54]. The Louvain algorithm maximizes modularity (a measure of community separation). Modularity rises from 0.0 as the number of intra-community edges increases relative to inter-community edges. Larger values of modularity reflect a more robust separation of the communities. The degree, average degree, and modularity class for each node were calculated by Gephi. Modularity resolution was set to 1.0 for the normalized unipartite graph and 1.15 for the normalized bipartite graph. For the normalized unipartite graph, two subjects were excluded from the final analysis as they formed communities with only one node. For the normalized unipartite graph, a cosine distance threshold of 0.4 was used to exclude weak edges. Feature means for each community were calculated by SPSS 28 (IBM, Chicago, IL). Differences between community feature means were tested by one-way ANOVA (SPSS). Feature maps were created with the *heat map* widget from Orange.

The concordance for set membership between communities was measured by the Jaccard Index [71] where J is the Jaccard Index and A and B are the set memberships of two communities:

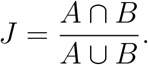

## RESULTS

### Bipartite network analysis

The normalized feature bipartite graph was partitioned into five communities (Figure 1a) with a modularity score of 0.247. Communities were named and color-coded by the one or two features with highest community means: fatigue (n=23), behavior (n=10), hypertonia/weakness (n=33), gait/sphincter (n=22), and sensory (n=25) (Figure 1b). ANOVA showed significant differences between communities for behavior (*p* < .001), cranial nerve (*p* = .008), eye movements (*p* < .001), fatigue (*p* < .001), gait (*p* = .029), hyperreflexia (*p* = .031), hypertonia (*p* < .001), incoordination (*p* = .002), sensory (*p* = .018), sphincter (*p* = .021), tremor (*p* = .006), and weakness (*p* = .001).

**Figure 1a.**
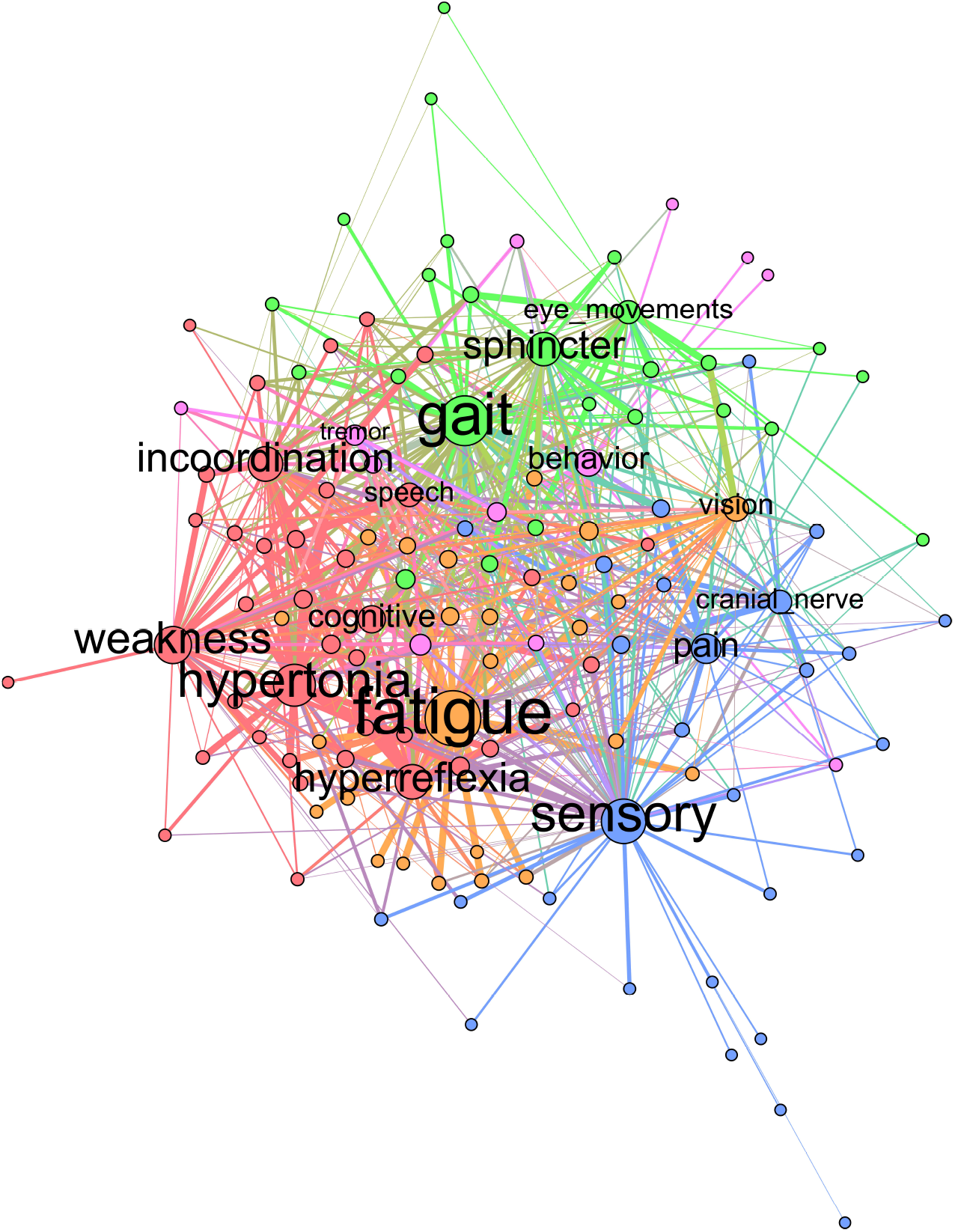
Bipartite graph of normalized features. Labeled nodes are features; unlabeled nodes are subjects. Weighted connections (edges) are between features and subjects.

**Figure 1b.**
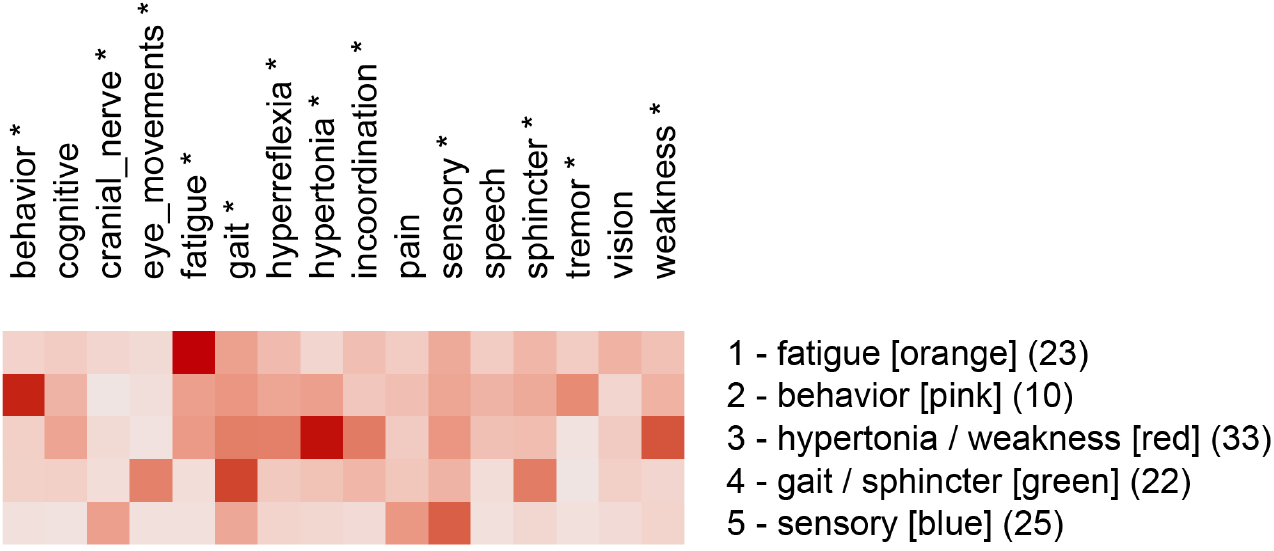
Feature map of the five communities identified by partitioning the bipartite graph with unnormalized features. Asterisks indicate features that differed significantly by community (One-way ANOVA, *p* < 0.05, *df* = 4)

### Unipartite network analysis

The normalized features unipartite graph was partitioned into five communities (Figure 2a) with a modularity score of 0.49. Communities were named by their predominant features: Community 1 (pain), Community 2 (fatigue), Community 3 (cognitive), Community 4 (sensory), and Community 5 (weakness/gait/hypertonia) (Figure 2b). ANOVA analysis showed significant differences between communities for behavior (*p* = .033), cognitive (*p* < .001), eye movements (*p* = .047), fatigue (*p* < .001), gait (*p* < .001), hyperreflexia (*p* = .014), hypertonia (*p* < .001), incoordination (*p* < .001), pain (*p* < .001), sensory (*p* < .001), and weakness (*p* = .037).

**Figure 2.** Modularity analysis of the bipartite graph of normalized data showed five communities. Community 1 was predominantly fatigue, Community 2 was predominantly behavioral, Community 3 was weakness and hypertonia, Community 4 was gait and sphincter, and Community 5 was predominantly sensory features.

**Figure 2a.**
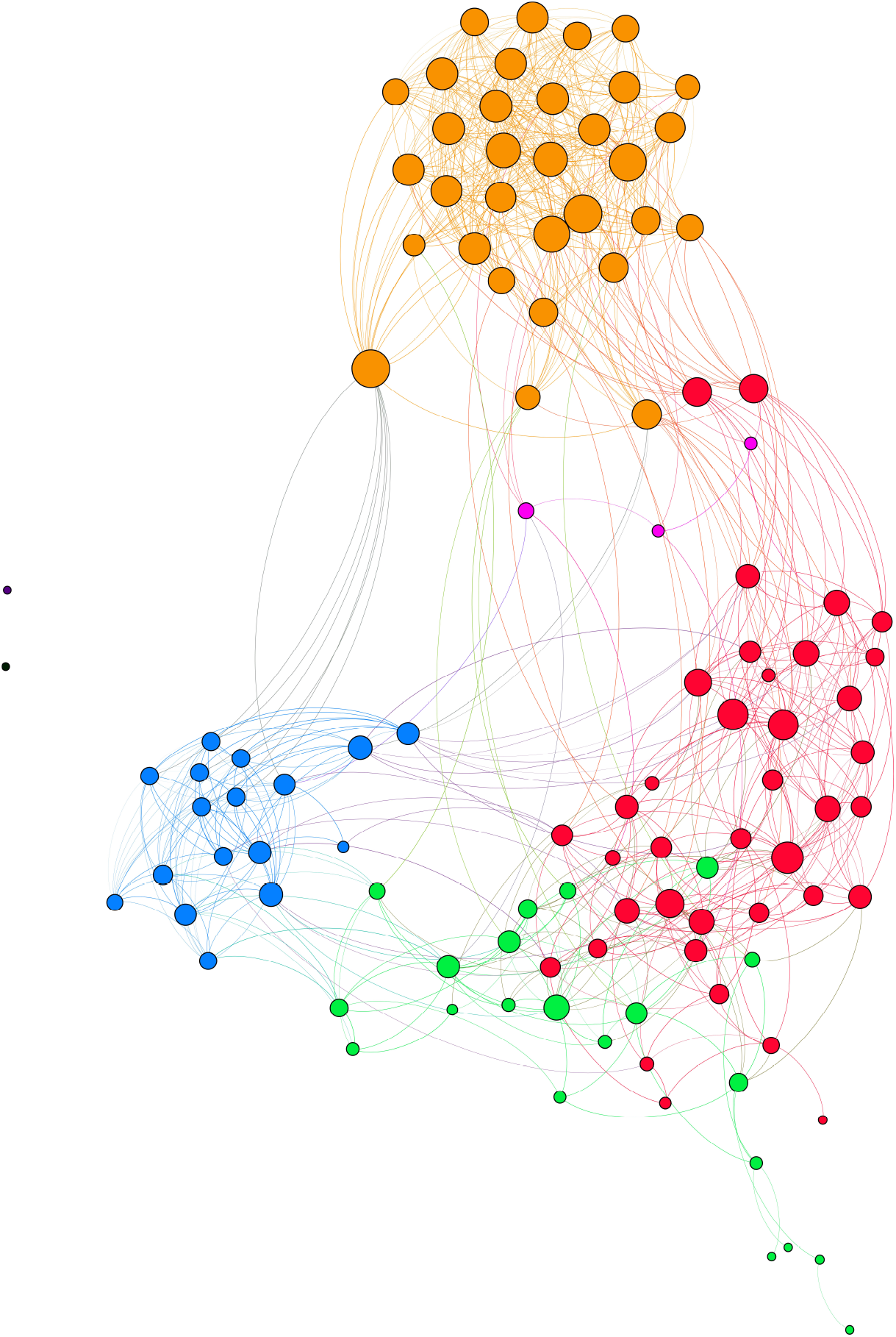
Unipartite graph based on normalized features. Nodes are subjects, and node size is proportional to the number of edges. The largest communities are gait/weakness/hypertonia (red, n=38) and fatigue (orange, n=32). Note the small cognitive community (pink, n=3).

**Figure 2b.**
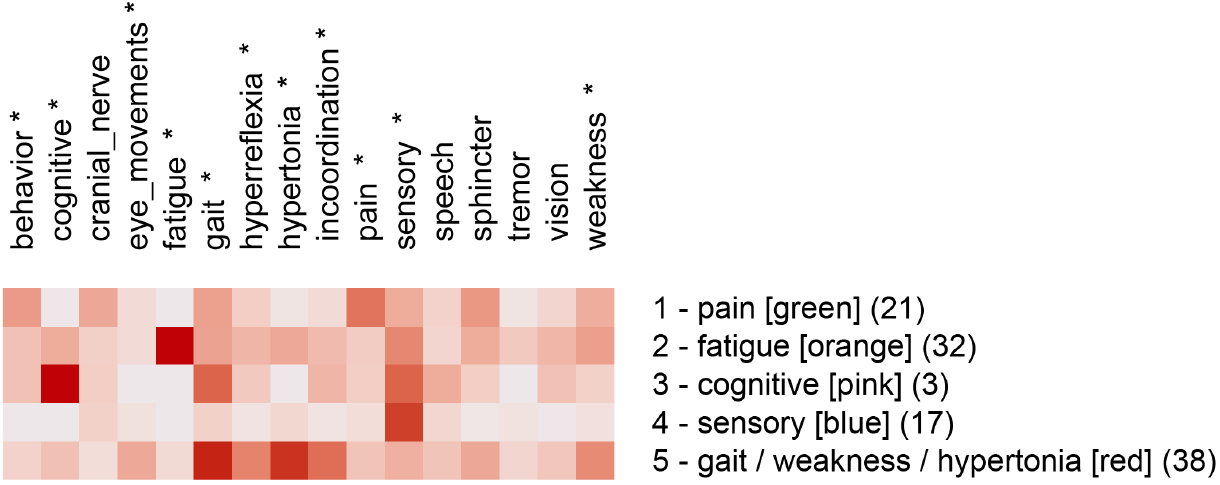
Feature map of the five communities identified by partitioning the unipartite network graph based on normalized features. Asterisks indicate features that differed significantly by community (One-way ANOVA, *p* < 0.05, *df* = 4)

### Community overlap across graphs

Although partitioning of the the bipartite and unipartite graphs produced somewhat different communities, similarities between community membership across graphs was notable. We used the Jaccard Index (a measure of set similarity) to assess similarity between communities. Membership for the *fatigue* and *sensory* communities was similar for the unipartite and bipartite graphs (Table 2). The unipartite graph community *gait/weakness/hypertonia* showed similarity to the bipartite graph communities *hypertonia/weakness* and *gait/sphincter*.

**Table 2.** Community concordances between bipartite and unipartite graphs as measured by Jaccard Index. Jaccard index is shown multiplied by 100. Fatigue and sensory communities show high concordance across graphs. The hypertonia/weakness and gait/sphincter communities from the bipartite graph overlap with the gait/weakness/hypertonia community from the unipartite graph.

|  | bipartite communities |  |  |  |  |
| --- | --- | --- | --- | --- | --- |
| unipartite communities | fatigue | behavior | hypertonia/weakness | gait/sphincter | sensory |
| pain | 0 | 24 | 6 | 10 | 21 |
| fatigue | 72 | 2 | 10 | 2 | 2 |
| cognitive | 0 | 0 | 9 | 9 | 9 |
| sensory | 0 | 0 | 2 | 3 | 56 |
| gait/weakness/hypertonia | 0 | 6 | 36 | 36 | 2 |

## DISCUSSION

Multiple sclerosis can present as sensory loss, weakness, incoordination, sphincter disturbance, diplopia, visual loss, cognitive impairment, fatigue, or even pain. We have used network analysis to explore whether we could identify distinct clinical subtypes of multiple sclerosis based on signs and symptoms. We first mapped the signs and symptoms of a cohort of multiple sclerosis subjects to concepts from a neuro-ontology. We then created a bipartite graph, where subjects and their signs and symptoms were nodes in a graph (Figure 1a). In a bipartite graph, subjects are connected to signs and symptoms and not to other subjects. When the signs and symptoms of a subject are converted to vectors, distances between subjects can be calculated so that subject nodes can be connected other subjects to form a unipartite graph (Figure 2a). Network analysis allowed us to identify communities of multiple sclerosis subjects who shared signs and symptoms in common. Partitioning of the unipartite and bipartite graphs based on modularity score identified communities with strong fatigue and sensory feature predominance. Both partitions had communities characterized by weakness that was combined with hypertonia or gait disorders. Partitioning of the bipartite graph produced a small community with behavioral changes (depression, anxiety, etc.) and a gait/sphincter community. Partitioning of the unipartite graph produced a small community with cognitive disorders and a medium-sized community with pain (Figure 2b.

Partitions of the unipartite graph yielded higher modularity scores than the bipartite graph, suggesting that the partition of the unipartite graph was more robust. The partition of the unipartite graph (Figure 2a) gave the highest modularity score and showed an especially robust separation of communities. The named communities for Figure 2b (*pain, fatigue, cognitive, sensory*, and *gait/weakness/hypertonia*) deserve special consideration as potentially identifiable multiple sclerosis subtypes. We found a strong overlap between the fatigue and sensory communities across both graphs (Table 2 as measured by the Jaccard Index. Significant overlap between the gait/weakness/hypertonia community from the unipartite graph with the gait/sphincter and hypertonia/weakness communities from the bipartite graph was noted. Although the partitioning of networks based on features suggests that identifiable MS subtypes may exist, variability across partitions does not permit a definitive characterization of subtypes at this time.

Although we did not correlate community features with MRI findings, the communities detected may reflect the anatomic location of MS lesions [72, 73]. While some identified communities may correlate with the anatomic localization of demyelinating plaques, correlations with other disease markers such as glial fibrillary acidic protein (GFAP) and neurofilament light chain (NFL) are possible.

Two strengths of this study should be mentioned. First, community detection was done by network analysis which offers an alternative to unsupervised machine learning algorithms based on cluster analysis. Second, we used subsumption and the hierarchical organization of signs and symptoms in an ontology to reduce the number of features used in the analysis [66]. The current study demonstrates that subsumption can successfully group signs and symptoms of MS subjects into superclasses. These superclasses can be used to characterize the clinical features of communities identified by network analysis.

The current study has several limitations. The sample size was small (N = 113). The small sample size raises questions of selection bias which may have influenced what communities of subjects were found by network analysis. Network analysis of larger sample sizes may detect more robust communities with a different profile of predominant features. Another limitation was that we evaluated only one partitioning algorithm (Louvain). Other partitioning algorithms are available and might yield different results. We used subsumption to reduce the number of clinical features from 244 to sixteen.

Different subsumption strategies would likely yield different results. We calculated distances between subjects using the cosine distance metric; other distance metrics are available and may have resulted in different results. The modularity scores of the partitions were modest (0.25 to 0.49), suggesting that the separation between communities was modest. Another limitation was that subjects in the study were diagnosed with the RRMS phenotype. Without further analysis, our data cannot be extrapolated to other disease course phenotypes. Our analysis did not consider the race or sex of the subjects, which could influence clinical subtype [8, 9, 10]. Finally, we partitioned MS subjects based on their accumulated signs and symptoms. Examining networks based on signs and symptoms at a single point in time would be instructive.

## CONCLUSIONS

MS phenotypes based on the clinical course are well-established and broadly accepted. Clinical subtypes of MS based on clinical presentation and clinical burden of signs and symptoms have long been recognized. Network analysis of signs and symptoms in MS subjects offers an avenue to identify clinical subtypes of MS. In particular, the feature maps (Figures 1b and 2b) suggest that identifiable subtypes of MS with predominant signs and symptoms related to weakness, sensory loss, behavioral changes, cognitive difficulties, pain, and fatigue deserve further investigation. Further investigation may reveal epigenetic, radiological, immunologic, or protein biomarkers that correlate with these MS clinical subtypes.

## Data Availability

All data produced in the present study are available upon reasonable request to the authors

## AUTHOR CONTRIBUTIONS

Design and concept by DBH and QHP. Data acquisition by MDC, DBH, QHP, and CO. Computations by DBH and QHP. Data analysis, data interpretation, and writing by DBH, QHP, MDC, CO, and DCWII. Revisions and approval by DBH, QHP, MDC, CO, and DCWII.

## DATA AVAILABILITY STATEMENT

Supporting data is available from the corresponding author on request.

